# Unmet demand, not reluctance: integrated HIV–tuberculosis community screening is highly acceptable in socioeconomically vulnerable adults in South India

**DOI:** 10.64898/2026.05.19.26353605

**Authors:** Meagan Karoly, Komal Jain, Madolyn Dauphinais, Senbagavalli Prakash Babu, Zeus Francis Kolengaden, Alessandra Christina Amaral Dutra, Rabina Bhandari, Bharath Lokireddy, Prakash Babu Narasimhan, C. Robert Horsburgh, Sonali Sarkar, Palanivel Chinnakali, Pranay Sinha

**Affiliations:** Boston Medical Center, Boston, MA, USA; Department of Preventive and Social Medicine, Jawaharlal Institute of Postgraduate Medical Education and Research (JIPMER), Puducherry, India; Qure.ai, Mumbai, India; University of Virginia School of Medicine, Charlottesville, VA, USA; Department of Immunology, Jawaharlal Institute of Postgraduate Medical Education and Research (JIPMER), Puducherry, India; Section of Infectious Diseases, Boston University Chobanian and Avedisian School of Medicine, Boston, MA, USA; Departments of Epidemiology, Global Health, and Biostatistics, Boston University School of Public Health, Boston, MA, USA

**Keywords:** tuberculosis, HIV, community screening, acceptability, active case finding, India

## Abstract

**Background:** Despite rising enthusiasm for active case-finding for TB, there have been concerns about conducting simultaneous HIV screenings due to perceived stigma, although the evidence to support this concern is scarce. We assessed the acceptability of integrated HIV–TB community screening and characterised participants’ motivations and prior testing history.

**Methods:** The SLIM study was a non-interventional cross-sectional study conducted in Puducherry (February 2023 – January 2024). In two community health camp-style screening events (one urban and one peri-urban), adults ≥18 years were offered TB screening via portable chest X-ray with AI-assisted interpretation (qXR, Qure.ai), plus sputum testing (Truenat), alongside point-of-care HIV testing. Structured questionnaires captured sociodemographics, prior testing history, and motivations for participation. Acceptability was pre-specified as >50% uptake.

**Results:** Of 273 eligible adults approached, 264 (96.7%) accepted integrated screening, nearly double our pre-specified threshold. Participants were predominantly low-income with limited formal employment. The dominant motivation was a desire to know one’s health status (HIV: 74.8%; TB: 73.7%), followed by convenience (16–17%). Prior HIV and TB testing was rare (7– 13% and 15–18%, respectively). Participation was uniformly high across demographic groups; however, the screened population skewed older and female (mean age 58 (standard deviation: 12.6) years; 67% female). Men under 45 comprised only 3.7% of participants — substantially below their 24.7% share in the Puducherry population per the most recent census.

**Conclusions:** Integrated HIV–TB screening achieved near-universal uptake in a socioeconomically vulnerable population with little prior testing exposure, contradicting concerns that community HIV screening would be poorly accepted in India. Integrated community-based screening should be scaled up as a cornerstone of TB elimination in high-burden settings. Crucially, because TB incidence in India peaks in the 15–45 age group and disproportionately affects men, targeted strategies to bring younger men and women into community screenings are essential.

## Dear Editor

India bears over a quarter of the global tuberculosis (TB) burden and is home to an estimated 2.56 million people living with HIV.^1,2,3^ Active case-finding (ACF) for TB has expanded substantially since the launch of India’s National Strategic Plan for TB Elimination in 2017, yet recent national assessments show that ACF quality indicators remain sub-optimal, with number-needed-to-screen values for population-based programmes well above acceptable thresholds.^4^ Concerns are even greater regarding the integration of HIV testing into community TB screening. Although HIV and TB are epidemiologically linked and the WHO recommends co-delivery wherever feasible, HIV testing in India remains largely facility-based and passive, with community-based approaches uncommon outside designated key populations, such as single male migrants and long distance truckers.^5^ Documented barriers to HIV testing uptake in India include HIV-related stigma, marginalised-group stigma, discrimination in public testing centres, and fear of adverse social consequences of a positive result.^6,7^ These concerns have made providers hesitant to offer community-based HIV screening — particularly when paired with other services — despite limited empirical evidence that general-population uptake is in fact low. Prior community HIV testing studies in India have largely been confined to antenatal integration8 or to key populations,^6^ leaving a gap in the evidence on acceptability among broader socioeconomically vulnerable adults.

We conducted a cross-sectional study of the acceptability and feasibility of integrated HIV–TB community screening at two community health camps in Puducherry, South India (July 2025 to October 2025). Camp 1 was situated at a non-communicable disease (NCD) outpatient clinic in a peri-urban area and Camp 2 at a school in central urban Puducherry. Both camps were held on Sundays when the venues were otherwise unoccupied. Adults (≥18 years) approached at camps were offered integrated TB and HIV screening alongside general health assessments. TB screening comprised portable chest radiography with AI-assisted interpretation (qXR, Qure.ai, Mumbai, India), with sputum Truenat MTB-RIF Dx testing (Molbio Diagnostics, Goa, India) for those with abnormal imaging. HIV screening used a point-of-care rapid diagnostic test. A structured questionnaire captured sociodemographics, modified Kuppuswamy socioeconomic scale, prior TB and HIV testing history, and self-reported motivations for accepting or declining either test. Acceptability was pre-specified as >50% uptake. All participants received a metal tiffin box (approximate value ₹150) as compensation, delivered regardless of whether they accepted screening. The study was approved by the institutional ethics committees of Boston Medical Center and JIPMER, Puducherry; all participants provided written informed consent in Tamil.

Of 273 eligible adults approached across both camps, 264 (96.7%) accepted integrated HIV–TB screening — nearly double our pre-specified threshold. Six participants (2.2%) declined both tests and three (1.1%) accepted TB testing only (Figure). No participant accepted HIV testing in isolation. Acceptance was similarly high at the peri-urban NCD clinic camp (95.5%) and the urban school camp (97.8%; p=1.00), though the two camps drew populations that differed in diabetes prevalence, occupational mix, and prior testing rates (Table S1). Participants were predominantly socioeconomically vulnerable: three-quarters (75.3%) reported per-capita household incomes in the lowest Kuppuswamy band (≤ ₹9,231/month), 55.1% of household heads were unemployed or retired, and 74.6% had middle-school education or below. Prior testing was strikingly uncommon: only 7–13% reported previous HIV testing and 15–18% prior TB testing, despite national ACF activities having been in place for over six years. The dominant self-reported motivation was a desire to know one’s health status (HIV, 74.8%; TB, 73.7%), followed by convenience (HIV, 17.0%; TB, 16.2%); altruistic or researcher-initiated motivations were uncommon. The screened population skewed older and female (mean age 57 years, SD 13; 67% female), with men under 45 comprising only 3.7% of participants — substantially below their roughly one-quarter share of the corresponding age–sex strata in the Puducherry census.^9^ Notably, among those who did present, acceptance was uniformly high across age and sex categories (100% among those <30 and ≥75; 96–97% in middle age bands; p = 0.49 comparing age of acceptors vs refusers).

**Figure 1:**
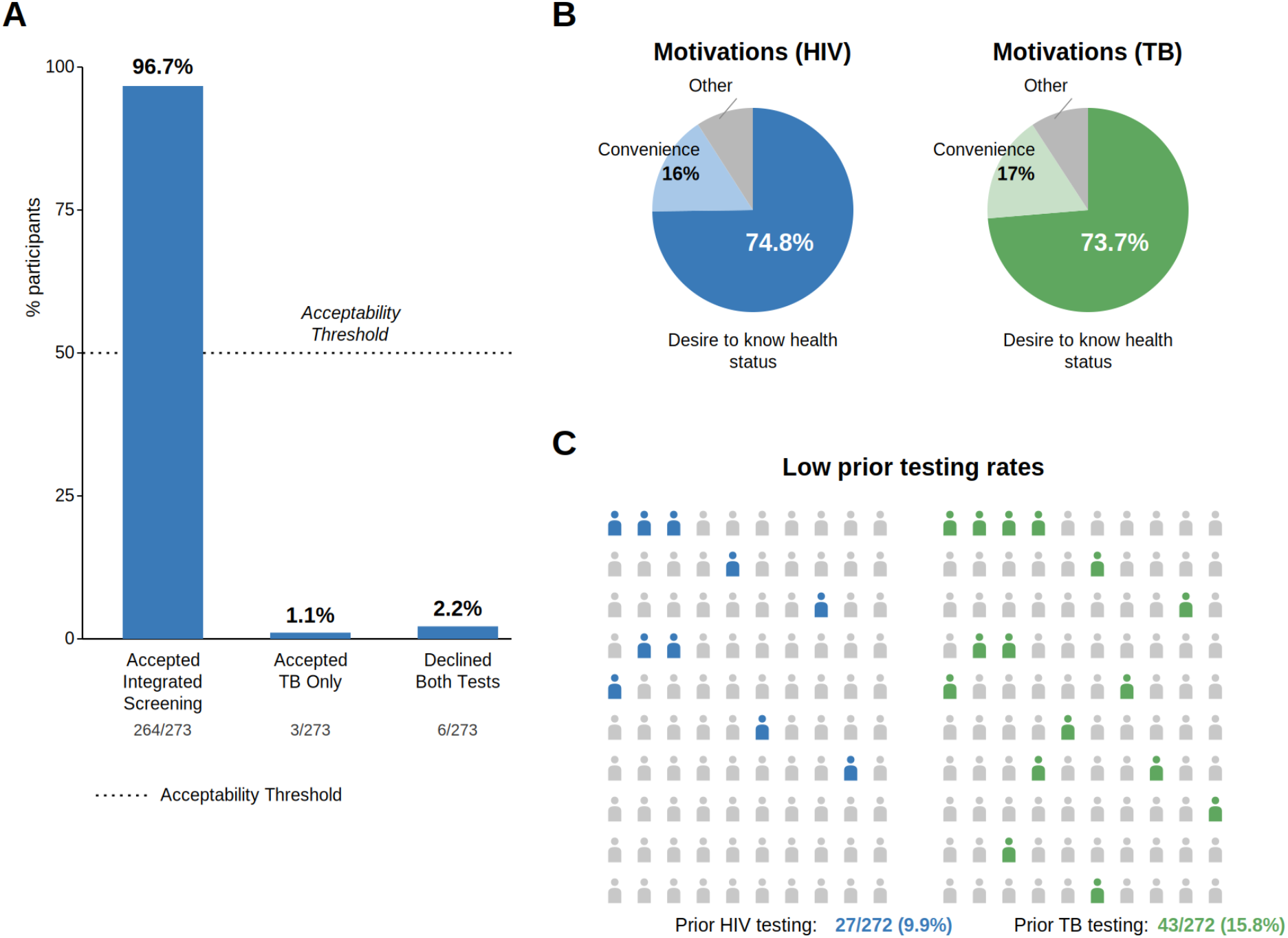
Integrated HIV–TB screening among 273 adults in Puducherry, South India: (A) acceptability, (B) self-reported motivations, and (C) prior testing rates.

These findings contradict the prevailing concern that community-based HIV screening in India would be poorly accepted due to stigma. In a socioeconomically vulnerable population with minimal prior testing exposure, integrated screening achieved near-universal10 uptake when offered alongside general health services. The distribution of motivations — overwhelmingly a desire to know one’s status, rather than altruism or accommodation of researchers — suggests that this uptake reflects genuine unmet demand rather than acquiescence. Combined with the strikingly low prior testing rates, these results point to an access barrier, not a motivation barrier. This reframes the implementation question from *“will communities accept integrated screening?”* to *“how do we bring integrated screening to communities who want it but cannot easily reach facility-based services?”* Scaled community-based delivery using portable, AI-supported tools offers one pragmatic answer, and could simultaneously advance TB elimination and the 95-95-95 HIV cascade targets.

The under-representation of younger men merits closer consideration, given that TB incidence in India peaks in the 15–45 age band and disproportionately affects males.^11^ During the urban school camp, our field social workers directly approached younger men in the surrounding neighbourhood; these invitations were largely declined, with potential participants citing weekend leisure at home. Field team members observed anecdotally that the tiffin-box incentive appeared more salient for older adults than for younger men, for whom nutrition-adjacent household items hold less appeal. While observational, these field notes align with our quantitative finding that younger men accepted screening readily when they did present but rarely presented in the first place — suggesting that camp-based models alone are unlikely to reach this demographic. Door-to-door outreach and age-appropriate incentive structures — such as cash incentives or raffles for consumer items — may be needed to close this gap.

Study limitations include the single-district setting, an acceptability denominator restricted to those who came to the health camps, a modest sample size, the potential influence of a participant incentive on camp attendance patterns, and the possibility that the in-person camp setting may have increased social desirability to accept screening. Nonetheless, our findings align with growing evidence that accessibility, rather than community-level stigma, may be the more important barrier to HIV testing uptake in general-population settings.^12^

In summary, integrated HIV–TB community screening was highly acceptable in a socioeconomically vulnerable South Indian population, suggesting that longstanding concerns about HIV-related stigma at the community level may be overstated for the general population. Scaling integrated community-based screening offers a pragmatic, high-yield pathway toward the dual goals of HIV and TB elimination.

## Data Availability

The de-identified data that support the findings of this study are available from the corresponding author upon reasonable request. Restrictions apply to the availability of these data, which contain potentially identifying information collected from a defined community population and were used under ethical approval that did not include public data sharing. Requests will be reviewed by the study investigators and the IRB to ensure participant confidentiality is protected.

## Supplementary Materials

**Table S1.** Sociodemographic characteristics, comorbidities, and HIV-TB screening acceptance by community screening camp venue, SLIM study, Puducherry, South India, 2025.

| Characteristic | Peri-urban NCD clinic (n=134) | Urban school (n=143) | Total (N=277) | p value |
| --- | --- | --- | --- | --- |
| **Demographics** |  |  |  |  |
| Age, years, median (IQR) | 59.0 (50.0–65.0) | 57.0 (49.0–65.0) | 58.5 (50.0–65.0) | 0.70 |
| Age group, n (%) |  |  |  | 0.16 |
| <30 | 2 (1.5) | 7 (4.9) | 9 (3.3) |  |
| 30–44 | 16 (12.0) | 15 (10.5) | 31 (11.2) |  |
| 45–59 | 49 (36.8) | 56 (39.2) | 105 (38.0) |  |
| 60–74 | 59 (44.4) | 50 (35.0) | 109 (39.5) |  |
| ≥75 | 7 (5.3) | 15 (10.5) | 22 (8.0) |  |
| Female sex, n (%) | 85 (63.9) | 100 (69.9) | 185 (67.0) | 0.31 |
| **Socioeconomic status** |  |  |  |  |
| Education of HoH, n (%) |  |  |  | 0.14 |
| No formal education | 33 (24.8) | 30 (21.6) | 63 (23.2) |  |
| Primary | 30 (22.6) | 37 (26.6) | 67 (24.6) |  |
| Middle | 44 (33.1) | 29 (20.9) | 73 (26.8) |  |
| High school | 19 (14.3) | 29 (20.9) | 48 (17.6) |  |
| Intermediate/diploma | 3 (2.3) | 5 (3.6) | 8 (2.9) |  |
| Graduate/professional | 4 (3.0) | 9 (6.5) | 13 (4.8) |  |
| Occupation of HoH, n (%) |  |  |  | <0.001 |
| Unemployed/retired | 74 (55.6) | 76 (54.7) | 150 (55.1) |  |
| Elementary occupation | 16 (12.0) | 18 (12.9) | 34 (12.5) |  |
| Skilled worker | 26 (19.5) | 16 (11.5) | 42 (15.4) |  |
| Skilled agriculture/fishery | 13 (9.8) | 6 (4.3) | 19 (7.0) |  |
| Clerk | 0 (0.0) | 17 (12.2) | 17 (6.2) |  |
| Other | 4 (3.0) | 6 (4.3) | 10 (3.7) |  |
| Lowest income band (≤₹9,231/mo), n (%) | 105 (79.5) | 99 (71.2) | 204 (75.3) | 0.12 |
| **Nutritional status** |  |  |  |  |
| BMI, kg/m<sup>2</sup>, median (IQR) | 24.9 (22.7–27.9) | 25.2 (22.7–29.6) | 25.1 (22.7–28.8) | 0.51 |
| BMI category (WHO cutoffs), n (%) |  |  |  | 0.59 |
| Underweight | 7 (5.2) | 9 (6.3) | 16 (5.8) |  |
| Normal | 61 (45.5) | 55 (38.7) | 116 (42.0) |  |
| Overweight | 43 (32.1) | 46 (32.4) | 89 (32.2) |  |
| Obese | 23 (17.2) | 32 (22.5) | 55 (19.9) |  |
| BMI category (Asian cutoffs), n (%) |  |  |  | 0.68 |
| Underweight (<18.5 kg/m<sup>2</sup>) | 7 (5.2) | 9 (6.3) | 16 (5.8) |  |
| Normal (18.5 – 22.9 kg/m<sup>2</sup>) | 29 (21.6) | 31 (21.8) | 60 (21.7) |  |
| Overweight (23 – 24.9 kg/m<sup>2</sup>) | 56 (41.8) | 50 (35.2) | 106 (38.4) |  |
| Obese (≥25 kg/m<sup>2</sup>) | 42 (31.3) | 52 (36.6) | 94 (34.1) |  |
| **Behavioural risk factors** |  |  |  |  |
| Smoking status, n (%) |  |  |  | 0.02 |
| Current | 1 (0.8) | 9 (6.5) | 10 (3.8) |  |
| Former | 4 (3.2) | 10 (7.2) | 14 (5.3) |  |
| Never | 121 (96.0) | 120 (86.3) | 241 (90.9) |  |
| Current alcohol use, n (%) | 15 (11.4) | 22 (15.8) | 37 (13.7) | 0.30 |
| **Self-reported comorbidities** |  |  |  |  |
| Diabetes mellitus, n (%) | 57 (44.9) | 31 (22.6) | 88 (33.3) | <0.001 |
| Asthma, n (%) | 7 (5.5) | 7 (5.1) | 14 (5.3) | 1.00 |
| Chronic bronchitis symptoms, n (%) | 1 (0.8) | 7 (5.1) | 8 (3.0) | 0.07 |
| Anaemia, n (%) | 4 (3.1) | 6 (4.4) | 10 (3.8) | 0.75 |
| **Clinical measurements** |  |  |  |  |
| Hypertension (SBP≥140 or DBP≥90), n (%) | 53 (39.6) | 51 (35.7) | 104 (37.5) | 0.54 |
| **Prior testing history** |  |  |  |  |
| Ever tested for TB, n (%) | 15 (11.3) | 28 (20.1) | 43 (15.8) | 0.05 |
| Ever tested for HIV, n (%) | 9 (6.8) | 18 (12.9) | 27 (9.9) | 0.11 |
| **HIV-TB Screening acceptance (n=273)** |  |  |  |  |
| Accepted both, n (%) | 128 (95.5) | 136 (97.8) | 264 (96.7) | 0.33 |
| Refused both, n (%) | 4 (3.0) | 2 (1.4) | 6 (2.2) | 0.44 |
| Accepted TB only, n (%) | 2 (1.5) | 1 (0.7) | 3 (1.1) | 0.62 |

Values are n (column %) for categorical variables or median (IQR) for continuous variables. Denominators for individual variables may vary owing to missing data; percentages are calculated among participants with non-missing responses.
Peri-urban NCD clinic = Camp 1, conducted at an unoccupied non-communicable disease outpatient clinic (July 2025). Urban school = Camp 2, conducted at an unoccupied school in central Puducherry (October 2025). Both camps were held on Sundays.
p values: Mann–Whitney U test for continuous variables; chi-square test or Fisher's exact test for categorical variables.
HoH = head of household; BMI = body mass index; SBP = systolic blood pressure; DBP = diastolic blood pressure; WHO = World Health Organization.
Chronic bronchitis symptoms defined as shortness of breath, wheezing, or coughing for $\geq 3$ months/year for $\geq 2$ consecutive years.
Screening acceptance denominators: peri-urban NCD clinic n=134, urban school n=139 (4 participants from the urban school camp had incomplete acceptance records and were excluded).

## References

1. World Health Organization. Global tuberculosis report 2023. Geneva: World Health Organization; 2023.

2. National AIDS Control Organisation, ICMR-National Institute for Research in Digital Health and Data Science. India HIV estimation 2025: technical report. New Delhi: Ministry of Health and Family Welfare, Government of India; 2025.

3. Joint United Nations Programme on HIV/AIDS. Country factsheet: India 2022. Geneva: UNAIDS; 2023.

4. Shewade HD, Kiruthika G, Ravichandran P, et al. Quality of active case-finding for tuberculosis in India: a national level secondary data analysis. Glob Health Action. 2023;16(1):2256129.

5. World Health Organization. Consolidated guidelines on HIV testing services. Geneva: World Health Organization; 2019.

6. Woodford MR, Chakrapani V, Newman PA, Shunmugam M. Barriers and facilitators to voluntary HIV testing uptake among communities at high risk of HIV exposure in Chennai, India. Glob Public Health. 2016;11(3):363–79.

7. Steward WT, Bharat S, Ramakrishna J, Heylen E, Ekstrand ML. Stigma is associated with delays in seeking care among HIV-infected people in India. J Int Assoc Provid AIDS Care. 2013;12(2):103–9.

8. Sinha A, Mugundu Ramachandran P, Kiran K, et al. Development and pilot testing of HIV screening program integration within public/primary health centres providing antenatal care services in Maharashtra, India. BMC Res Notes. 2014;7:177.

9. Office of the Registrar General and Census Commissioner, India. District census handbook: Puducherry (Series 35, Part XII-A). New Delhi: Ministry of Home Affairs; 2014.

10. Laxmeshwar C, Hegde A, Dange A, Mariyappan K, Soosai M, Mane S, et al. Acceptability, usability, and willingness to pay for HIV self-test kits distributed through community-based, PLHIV network-led and private practitioners models in India: results from the STAR III Initiative. J Int AIDS Soc. 2024;27:e26348.

11. Ministry of Health and Family Welfare, Government of India. India TB report 2023. New Delhi: Central TB Division; 2023.

12. Mamulwar M, Prasad VS, Nirmalkar A, et al. Community-based point-of-care testing to identify new HIV infections: a cross-sectional study from Pune, India. Medicine (Baltimore). 2021;100(46):e27817.

